# Cardiovascular Health at Midlife and Alzheimer Disease Biomarkers

**DOI:** 10.64898/2026.04.15.26350968

**Authors:** Christina S. Dintica, Xiaqing Jiang, Leslie M. Shaw, R. Nick Bryan, Kristine Yaffe

## Abstract

**Background:** Cardiovascular health factors are associated with cognitive decline and risk of dementia, including Alzheimer disease (AD); however, this has been mostly studied in late life. We investigated whether vascular and lifestyle factors are associated with AD plasma and imaging biomarkers in midlife.

**Methods:** We investigated 1,406 participants from the Coronary Artery Risk Development in Young Adults (CARDIA) study with information on vascular and lifestyle factors framed from the American Heart Association (AHA) “life’s essential 8” (LE8) guidelines for cardiovascular health at early midlife (mean age 45.0 ± SD 3.6) and AD biomarkers in late midlife (mean age 60 ± SD 3.5). LE8 was calculated and categorized into poor (0-49), intermediate (50-79), and ideal (80-100) cardiovascular health, based on 8 components including smoking, diet, body mass index (BMI), sleep, fasting glucose, blood pressure, cholesterol, and physical activity. We assessed the AD plasma biomarkers phosphorylated tau 217 (ptau-217) and amyloid beta 42/40 ratio (Aβ42/40) and the Spatial Pattern of Abnormality for Recognition of Early AD (SPARE-AD), an algorithm that characterizes AD-like brain atrophy on brain MRI. We used linear regression to examine the association between LE8 and log transformed and standardized AD biomarker measures adjusting for age, sex, race, education, and kidney function.

**Results:** Compared to ideal LE8, intermediate (67.9% of participants) and poor (12.6%) LE8 was associated with lower Aβ42/40 (adjusted mean difference: −2.37, 95% CI: −2.38 to −2.36 and −2.38, 95% CI: −2.40 to −2.36, respectively). There was no association between the LE8 group and ptau-217 level. Moreover, compared to ideal LE8 participants, those with poor LE8 had higher SPARE-AD atrophy pattern (adjusted mean difference: −0.71, 95% CI: −0.81 to −0.62).

**Conclusion:** These findings indicate that poor cardiovascular health in midlife, as defined by the AHA LE8, is linked to less favorable early AD biomarker profiles, particularly reflecting greater amyloid burden and structural brain changes.

## INTRODUCTION

Alzheimer’s disease (AD) is a progressive neurodegenerative disorder with a prolonged preclinical phase during which neuropathological changes accumulate in the absence of overt symptoms. ^1^ Increasing evidence suggests that midlife may represent a critical window for the initiation of these pathological processes, underscoring the need to identify modifiable risk factors during this period.^2^ Cardiovascular health has emerged as one important factor, with a growing body of literature linking vascular risk to late-life cognitive decline and dementia including vascular dementia and AD.^3^ Vascular risk factors such as hypertension, diabetes, and dyslipidemia not only increase the overall likelihood of developing AD^4^ and in some studies correlate with enhanced β-amyloid accumulation in cognitively normal older individuals.^5,6^ Most evidence to date is from older adults, leaving a gap in understanding how cardiovascular health in midlife relates to early biomarkers of AD. Yet, if there is an association at midlife, this would support that this life phase would be an ideal time for primary prevention.

To address this knowledge gap, we examined the association between the American Heart Association (AHA) recently updated definition of ideal cardiovascular health through the “Life’s Essential 8” (LE8) framework and AD-related biomarkers in a diverse midlife cohort. Specifically, we evaluated associations with plasma-based biomarkers of amyloid (Aβ42/40), tau (ptau-217), as well as an MRI imaging-based marker of AD-like brain atrophy.^10^ Understanding how cardiovascular health in midlife relates to early indicators of AD may inform preventive strategies and promote brain health across the life course.

## METHODS

### Data availability

Requests for access to the data for this study can be made at the CARDIA website: https://www.cardia.dopm.uab.edu/.

### Study population

The Coronary Artery Risk Development in Young Adults (CARDIA) Study is a large prospective cohort study investigating the development of and risk factors for cardiovascular disease.^9^ Briefly, starting in 1985, 5115 Black and White community-dwelling adults between 18 and 30 years of age were recruited from community-based samples of four US cities (Birmingham, Alabama; Chicago, Illinois; Minneapolis, Minnesota; and Oakland, California). Within each center, recruitment was balanced by sex, age, and educational level. At each examination, participants provided written informed consent, and study protocols were reviewed by institutional review boards at each study site and the CARDIA Coordinating Center; starting in 2020, a single institutional review board at the University of Alabama, Birmingham provided this review.^9^

A total of 3,526 non-demented participants attended the CARDIA Year 20 examination (2005– 2006), at which time all LE8 components were assessed; this served as the baseline for the present study. Plasma biomarker data were subsequently collected 15 years later at the Year 35 examination (2020–2021) in 1,406 participants. Of these, a subsample of 601 participants also underwent MRI at Year 35.

### Life Essential 8 Cardiovascular Health Score

The updated AHA LE8 metric includes healthy diet, participation in physical activity, avoidance of nicotine, healthy sleep, healthy weight, and healthy levels of blood lipids, blood glucose, and blood pressure.^7^ We used the Mediterranean diet (MedDiet) as a measure of diet quality. Physical activity was measured with the CARDIA Physical Activity History questionnaire, which queries the amount of time per week spent in 13 categories of leisure, occupational, and household physical activities over the past 12 months summarized as units of total activity incorporating moderate and high intensity activities. Nicotine use was assessed via self-reported question “Have you ever used any tobacco product such as cigarettes, cigars, tobacco pipe, chewing tobacco, snuff, nicotine chewing gum, or a nicotine patch?”. If participants answered yes, the answers were further coded into current use, or the most recent past use. Hours of sleep were also ascertained by self-report. Body mass index (BMI) was calculated as weight in kilograms divided by height in meters squared. Plasma total and high-density lipoprotein (HDL) cholesterol was measured enzymatically as previously described,^13^ and used to estimate non-HDL cholesterol. A digital blood pressure monitor (Omron HEM-907XL; Online Fitness) was used to measure systolic and diastolic blood pressure. Glucose levels were determined through fasting glucose and/or HbA1c.

The detailed definitions and scoring for the component metrics of LE8 are provided in Supplementary Table 1. Each component was scored from 0 to 100 points, with higher scores indicating healthier CVH.^14^ The overall LE8 score was calculated as the mean of the 8 metrics, ranging from 0 (lowest) to 100 (highest), and the score was categorized as poor LE8 for scores ranging from 0 to 49, intermediate for scores ranging from 50 to 79, and ideal for scores ranging from 80 to 100 as suggested by the AHA and previous literature.^7,14–17^

### Blood Processing and Plasma Biomarkers

Blood samples were drawn at the Year 35 clinic visit, processed and stored within 90 min (stick-to-freezer) at −70°C until assayed between March and May 2024 at the Laboratory of Pathology and Laboratory Medicine at the Hospital of the University of Pennsylvania (Director, Dr.Shaw). Plasma Aβ1-42, Aβ1-40, and p-tau217 were quantified using the Fujirebio fully automated Lumipulse chemiluminescence enzyme immunoassay platform (Lumipulse G1200, Fujirebio Diagnostics) with results reported in pg/mL within the manufacturer’s specified ranges.

To ensure quality assurance, calibration standards and manufacturer-provided aqueous quality controls (QCs) were included in each analytical run. In addition, six plasma pool QC samples derived from individuals with a range of biomarker profiles were analyzed in parallel. Across 15 analytical runs, the average concentrations and coefficients of variation (%CV) for p-tau217, Aβ1-42, and Aβ1-40 were consistently within acceptable limits (e.g., Con1: p-tau217 = 0.452 pg/mL, %CV = 3.76; Aβ1-42 = 20.34 pg/mL, %CV = 3.84; Aβ1-40 = 212.88 pg/mL, %CV = 2.87). Full QC performance data are summarized in Supplementary Table 2.

### Imaging data

Also at the Year 35 visit, a subset of CARDIA participants completed MRI brain scanning at each of the four clinic sites with approximate balance within four strata of race (Black, White) and sex (male/female).^13^ We used a composite brain atrophy biomarker, SPARE-AD (Spatial Pattern of Abnormality for Recognition of Early Alzheimer’s disease) index^10^, derived from volumetric measures of structural MRI data. The SPARE-AD index is a neuroimaging biomarker tool developed to identify early stage of AD by capturing spatial patterns of brain atrophy associated with the disease. This index has been widely used in AD-related studies and has shown great performance in predicting AD risk.^18,19^ More positive SPARE-AD index indicates a higher AD risk, while more negative values imply lower AD risk.

### Covariates

Demographic characteristics at our analytic baseline (Year 20) including age, sex, education, and race were based on self-report. Apolipoprotein E (*APOE*) phenotype was determined and participants were categorized as having any ε4 versus no ε4 allele. ^20^ Estimated glomerular filtration rate (eGFR) was calculated using the 2021 CKD-EPI creatinine equation without race, which incorporates serum creatinine, age, and sex to estimate kidney function in mL/min/1.73 m^2^.^21^

### Statistical Analysis

Participant characteristics across the three LE8 categories were compared using χ^2^ tests for categorical variables and one-way ANOVA for continuous variables. For biomarker analyses, we assessed linear trends across LE8 categories by modeling the ordinal category variable as a continuous term and reporting the corresponding p-trend. The blood biomarkers were log transformed to handle skewness and standardized to allow for comparison between the biomarkers.

We used linear regression models with robust standard errors and margins post estimation to derive adjusted mean difference in biomarkers among participants in the LE8 groups in the overall sample, adjusted for age, sex, race, and education. Models were further adjusted for *APOE* ε4 status, given its strong influence on amyloid and tau pathology and its potential to confound associations between cardiovascular health and AD biomarkers.^23,24^

Moreover, we used linear regression models with robust standard errors and margins post estimation to derive difference in SPARE-AD among participants the LE8 groups in the MRI sample, adjusted for age, sex, race, and education.

Linear regression models were also used to estimate the association between the individual LE8 components (poor vs. ideal/intermediate) and the AD biomarkers. We adjusted models for age, race, sex, education, and eGFR; eGFR was included in the models as previous findings demonstrated an association with increased AD biomarker levels.^22^

Moreover, we tested interactions between LE8 group by sex, race and *APOE* ε4 status on the plasma biomarkers and the SPARE-AD index.

All statistical analyses were conducted using STATA version 15.1 and R-Studio (Version 4.0.4).

## RESULTS

In the main sample, 267 (19.5%) participants had ideal LE8, 961 (67.9%) had intermediate, and 178 (12.6%) had poor LE8. Participants with poor LE8 were more likely to be female, Black, have fewer years of education, and be *APOE* ε4 carriers. There was a significant linear trend across LE8 categories for Aβ42/40 (*p*-trend <0.001), with intermediate and poor LE8 associated with lower Aβ42/40 (adjusted mean difference: −0.03, 95% CI: −0.09 to −0.04 for intermediate; −0.19, 95% CI: −0.33 to −0.05 for poor) compared to ideal LE8 (0.21, 95% CI: 0.09 to 0.33). No significant differences in ptau-217 were observed across LE8 categories (Table 2). There was no interaction between LE8 group by sex, race or *APOE* ε4 status on the plasma biomarkers.

**Table 1.** Study population characteristics by baseline by Life’s Essential 8 CVH score (n=1,406).

| Variable | **Ideal (>80)** | **Intermediate (50-80)** | **Poor (<50)** | *p* value |
| --- | --- | --- | --- | --- |
| Mean (sd) or N (%) | n= 267 (19.5%) | n= 961 (67.9%) | n= 178 (12.6%) |  |
| Age | 45.46 (3.4) | 45.2 (3.6) | 45.0 (3.6) | 0.390 |
| Female sex | 182 (65.9) | 515 (53.6) | 107 (60.1) | 0.001 |
| Education, years | 14.4 (1.9) | 14.2 (2.2) | 15.2 (2.1) | <0.001 |
| Black Race | 53 (19.2) | 444 (46.2) | 121 (68.0) | <0.001 |
| LE8 components |  |  |  |  |
| Current smoking | 9 (3.3) | 160 (16.9) | 61 (34.5) | <0.001 |
| MedDiet score | 32.6 (5.8) | 25.6 (10.0) | 20.2 (11.1) | <0.001 |
| Body mass index<br>(kg/m<sup>2</sup>) | 24.0 (3.2) | 29.4 (7.1) | 35.8 (8.1) | <0.001 |
| Hours of<br>sleep/night | 7.0 (1.0) | 6.9 (3.3) | 6.3 (3.5) | 0.027 |
| Fasting glucose,<br>mg/dL | 89.5 (7.6) | 96.5 (17.0) | 108.1 (33.7) | <0.001 |
| Systolic blood<br>pressure, mm Hg | 107.3 (9.1) | 115.6 (13.8) | 125.2 (17.4) | <0.001 |
| Non-HDL, mg/dL | 110.2 (26.2) | 132.8 (33.7) | 149.8 (42.2) | <0.001 |
| Physical activity | 480.1 (266.1) | 343.5 (281.2) | 172.5 (194.8) | <0.001 |
| Apolipoprotein e4 | 66 (25.6) | 278 (31.8) | 61 (38.1) | 0.024 |

391 HDL= high-density lipoprotein.

**Table 2.**
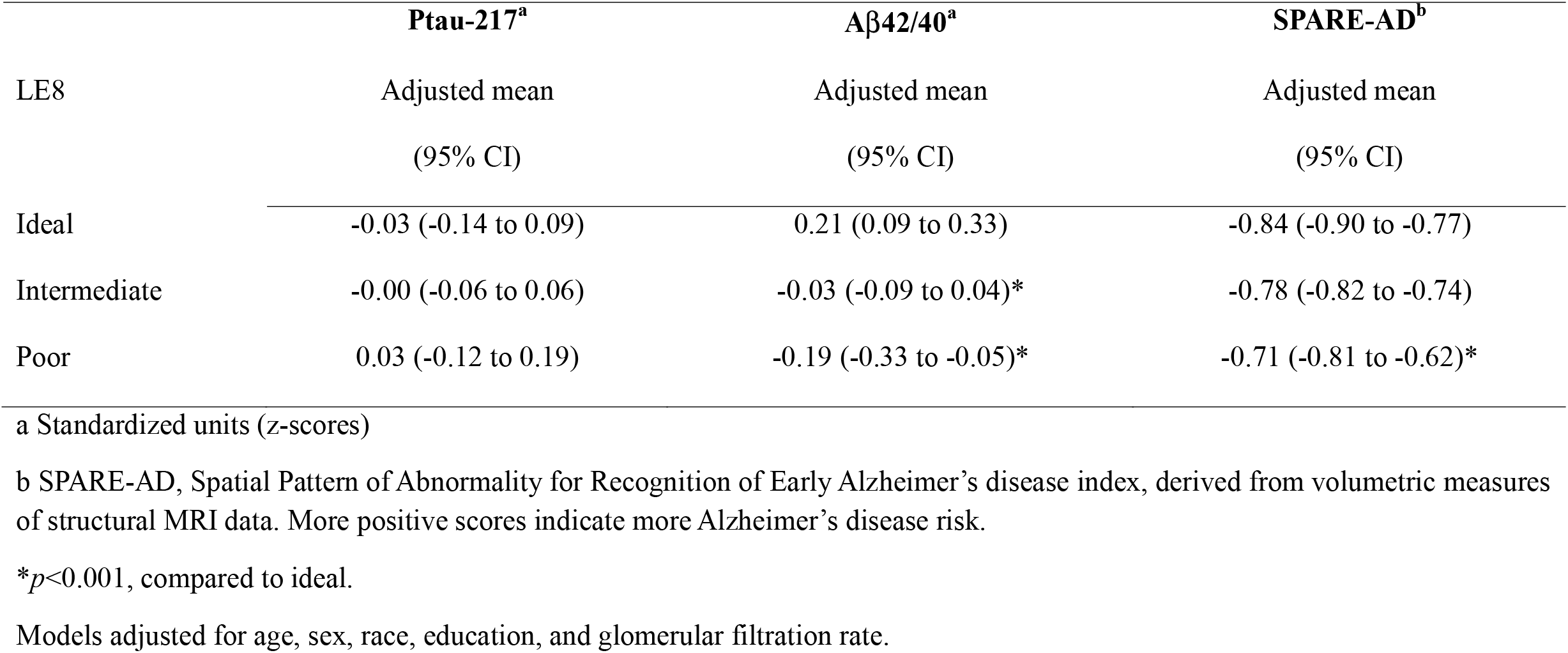
Association between the Life’s essential 8 and AD biomarkers in midlife non-demented adults (n=1,406).

|  | **Ptau-217<sup>a</sup>** | **Aβ42/40<sup>a</sup>** | **SPARE-AD<sup>b</sup>** |
| --- | --- | --- | --- |
| LE8 | Adjusted mean<br>(95% CI) | Adjusted mean<br>(95% CI) | Adjusted mean<br>(95% CI) |
| Ideal | -0.03 (-0.14 to 0.09) | 0.21 (0.09 to 0.33) | -0.84 (-0.90 to -0.77) |
| Intermediate | -0.00 (-0.06 to 0.06) | -0.03 (-0.09 to 0.04)* | -0.78 (-0.82 to -0.74) |
| Poor | 0.03 (-0.12 to 0.19) | -0.19 (-0.33 to -0.05)* | -0.71 (-0.81 to -0.62)* |

a Standardized units (z-scores)
b SPARE-AD, Spatial Pattern of Abnormality for Recognition of Early Alzheimer's disease index, derived from volumetric measures of structural MRI data. More positive scores indicate more Alzheimer's disease risk.
\* $p < 0.001$ , compared to ideal.

For the individual LE8 components, higher blood pressure was associated with elevated p-tau217 levels. For Aβ42/40, higher BMI and glucose and worse sleep and diet were associated with lower ratios (reflecting greater amyloid pathology) (Figure 1).

**Figure 1.**
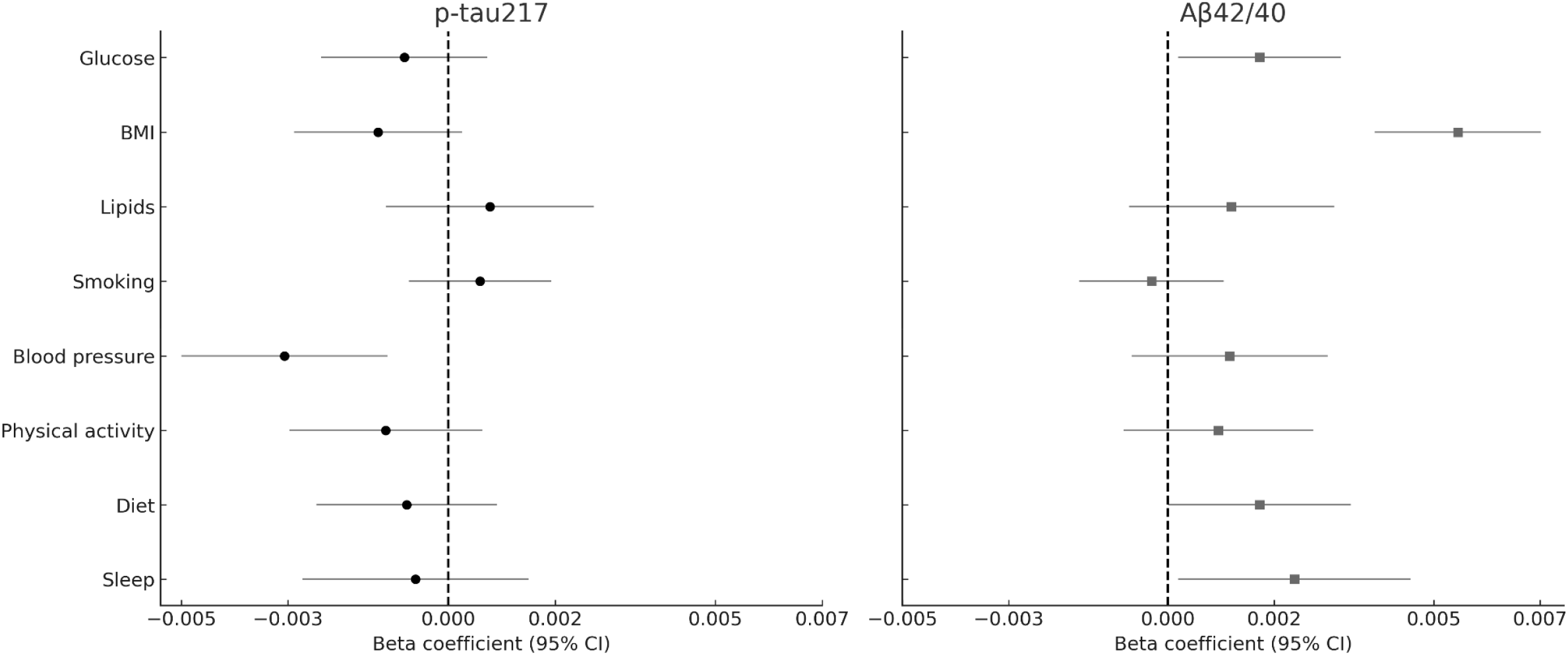
Association between the Life’s essential 8 components and AD biomarkers (n=1,406). Models adjusted for age, sex, race, education, and glomerular filtration rate.

In the MRI sample, 122 (20%) had ideal LE8, 420 (69%) had intermediate, and 67 (11%) had poor LE8. Participants with poor LE8 had higher SPARE-AD atrophy pattern scores (adjusted mean difference: −0.71, 95% CI: −0.81 to −0.62) compared to ideal LE8 (−0.84, 95% CI: −0.90 to −0.77), representing approximately a 15% greater burden of AD-like brain atrophy. A significant p-for trend was observed across categories (*p*<0.001). Among the individual LE8 components, higher non-HDL cholesterol was associated with greater SPARE-AD (β = −0.001, 95% CI: −0.003 to −0.001). There was no evidence of effect modification by sex, race, or *APOE* ε4 status.

## DISCUSSION

In this large, community-based cohort of middle-aged adults, we found that better cardiovascular health, as defined by the American Heart Association’s Life’s Essential 8 (LE8) guidelines, was associated with more favorable Alzheimer’s disease (AD) biomarker profiles. Specifically, participants with ideal cardiovascular health in early midlife had significantly higher plasma Aβ42/40 ratio and lower levels of AD pattern brain atrophy compared to those with intermediate or poor LE8 scores. These associations remained after adjusting for key demographic and clinical covariates, including *APOE* ε4 status. In contrast, no significant associations were observed between LE8 categories and ptau-217. However, several individual LE8 components, including lower blood pressure and fasting glucose, were associated with lower ptau-217 levels. Taken together, these findings suggest that maintaining cardiovascular health in midlife may have neuroprotective effects and support early intervention strategies targeting vascular risk to promote brain health. To our knowledge, this is one of the first studies to evaluate the updated LE8 metric in relation to AD biomarkers within a midlife, community-based cohort, providing novel insights into the vascular contributions to early AD-related changes.

These results complement those linking cardiovascular risk to late-life cognitive decline and dementia by showing associations between midlife cardiovascular health and early indicators of AD pathophysiology. The observed association between higher LE8 scores and increased Aβ42/40 ratios is particularly notable, as lower Aβ42/40 is thought to reflect greater brain amyloid deposition, an early hallmark of AD. These differences correspond to approximately a 2–3% lower Aβ42/40 ratio, a magnitude consistent with early amyloid changes observed in preclinical AD studies.^25^ This result aligns with prior studies suggesting that vascular health is associated with amyloid burden, including positron emission tomography (PET) imaging studies and CSF biomarker investigations in older adults.^26–28^ Our findings provide novel evidence that these associations may be detectable as early as midlife using plasma-based biomarkers, thereby supporting the utility of LE8 as a risk stratification tool well before clinical symptoms arise.

Potential mechanisms linking cardiovascular health to amyloid pathology include impaired clearance of amyloid-β via perivascular drainage pathways, blood–brain barrier dysfunction, and chronic cerebrovascular insufficiency. These processes can all be exacerbated by vascular risk factors such as hypertension, hyperlipidemia, and diabetes. Impaired perivascular clearance is increasingly recognized as a contributor to amyloid accumulation,^29^ while BBB breakdown has been observed in early AD stages, even before neurodegeneration or atrophy.^30^ Moreover, hypertension compromises vascular integrity and brain perfusion, facilitating amyloid deposition and cerebrovascular pathology.^31^

Interestingly, we did not observe statistically associations between summary LE8 and ptau-217, despite prior work implicating vascular risk factors in tau pathology and neurodegeneration.^32,33^ One potential explanation is that amyloid pathology precedes tau changes in the AD cascade, and thus, midlife may be too early to observe robust associations with downstream biomarkers such as ptau-217.^34^ Alternatively, our use of a relatively healthy, middle-aged cohort may have limited the range of biomarker expression, thereby attenuating effect sizes. However, our analysis of individual LE8 components revealed that specific aspects of cardiovascular health—notably, blood pressure and fasting glucose—were independently associated with lower ptau-217 levels. These findings underscore the utility of the LE8 as a comprehensive measure of cardiovascular health linked to early AD biomarkers, while also highlighting that examining its individual components can inform more tailored intervention strategies.

The association between LE8 and structural brain changes, as captured by the SPARE-AD index, further supports the hypothesis that cardiovascular health is linked to neurodegeneration. Participants with poor LE8 scores exhibited significantly higher SPARE-AD values, indicating greater AD-like atrophy. This finding aligns with previous imaging studies that have shown vascular risk factors, including hypertension, diabetes, and obesity, are associated with regional brain atrophy and reduced gray matter volume.^1^

Our results have several implications for public health and clinical practice. First, they reinforce the importance of cardiovascular health in midlife as a modifiable determinant of brain aging and AD risk. By demonstrating associations between LE8 and early biomarkers of AD, our study supports the inclusion of brain health as a key outcome in cardiovascular prevention efforts.

Second, our findings highlight the utility of the updated LE8 framework, which incorporates newer dimensions of health such as sleep and non-smoking status, in capturing relevant risk profiles. Third, the use of plasma biomarkers and neuroimaging indices in a diverse cohort underscores the feasibility of implementing such markers in large-scale population health monitoring and intervention trials.

Strengths of our study include the large sample size, well-characterized cohort with longitudinal follow-up, comprehensive assessment of cardiovascular health, and inclusion of both blood-based and imaging biomarkers. The use of the LE8 framework provides a contemporary and standardized measure of cardiovascular health that aligns with national prevention goals.

Several limitations should be noted. Although plasma biomarkers offer a minimally invasive and scalable approach, they may be less sensitive than CSF or imaging-based modalities in detecting early AD pathology. The composite LE8 score may mask differential effects of individual components; future studies should explore alternative weighting schemes or clustering approaches to refine risk prediction. Finally, the MRI subsample was smaller and power to detect differences was lower. As this was an observational study, the findings demonstrate associations rather than causal relationships, and longitudinal studies are needed to confirm these relationships over time.

In conclusion, our findings suggest that poor cardiovascular health in midlife, as captured by the AHA LE8, is associated with more unfavorable AD biomarker profiles, particularly regarding amyloid pathology and structural brain changes. These results support the integration of cardiovascular and brain health promotion efforts and emphasize the importance of early-life prevention strategies to reduce the burden of AD and related dementias. Continued longitudinal investigation of these relationships will be critical to further elucidate the pathways linking cardiovascular and neurodegenerative health across the life course.

## Data Availability

The CARDIA dataset can be requested via the study website https://www.cardia.dopm.uab.edu/. CARDIA study data are available to affiliated and non-affiliated investigators. See the study website for further details: http://www.cardia.dopm.uab.edu/invitation-to-new-investigators.

## ACKNOWLEDGEMENTS

The Coronary Artery Risk Development in Young Adults Study (CARDIA) is supported by contracts 75N92023D00002, 75N92023D00003, 75N92023D00004, 75N92023D00005, and 75N92023D00006 from the National Heart, Lung, and Blood Institute (NHLBI).

CARDIA was also partially supported by the Intramural Research Program of the National Institute on Aging (NIA) and an intra-agency agreement between NIA and NHLBI (AG0005).

