## Supplementary Table 1 for "Cardiovascular Health at Midlife and Alzheimer Disease Biomarkers"

**Supplementary Table 1:** Scoring Scheme for the Life’s Essential 8 Score Components.

| **Health Factors** | | **Health Behaviors** | |
| --- | --- | --- | --- |
| **Blood Pressure*** | **Points** | **Diet**  **Quantile of the** **Mediterranean diet** **(MedDiet)** | **Points** |
| Systolic BP <120 mmHg and  Diastolic BP <80 mmHg | 100 | ≥95^th^ percentile | 100 |
| Systolic BP 120-129 mmHg and  Diastolic BP <80 mmHg | 75 | 75^th^-94^th^ percentile | 80 |
| Systolic BP 130-139 mmHg or  Diastolic BP 80-89 mmHg | 50 | 50^th^-74^th^ percentile | 50 |
| Systolic BP 140-159 mmHg or  Diastolic BP 90-99 mmHg | 25 | 25^th^-49^th^ percentile | 25 |
| Systolic BP ≥160 mmHg or  Diastolic BP ≥100 mmHg | 0 | 1^st^-24^th^ percentile | 0 |
| **Non-HDL Cholesterol*** |  | **Physical Activity**  **Self-Reported Minutes of Moderate or Vigorous Intensity Activity per Week** |  |
| <130 mg/dL | 100 | ≥150 minutes | 100 |
| 130-159 mg/dL | 60 | 120-149 minutes | 90 |
| 160-189 mg/dL | 40 | 90-119 minutes | 80 |
| 190-219 mg/dL | 20 | 60-89 minutes | 60 |
| ≥220 mg/dL | 0 | 30-59 minutes | 40 |
| **Blood Glucose**  **Glycosylated Hemoglobin A_1_C** |  | 1-29 minutes | 20 |
| Fasting blood glucose <100 mg/dL or HbA_1_C <5.7 gm% with no history of diabetes mellitus | 100 | 0 minutes | 0 |
| Fasting blood glucose 100-125 mg/dL or HbA_1_C 5.7-6.5 gm% with no history of diabetes mellitus | 60 | **Smoking**  **Self-Reported Cigarette Smoking** |  |
| HbA_1_C <7.0 gm% with diabetes | 40 | Never smoked | 100 |
| HbA_1_C 7.0-7.9 gm% | 30 | Former smoker, quit ≥5 years ago | 75 |
| HbA_1_C 8.0-8.9 gm% | 20 | Former smoker, quit 1-<5 years ago | 50 |
| HbA_1_C 9.0-9.9 gm% | 10 | Former smoker, quit <1 year ago  Currently using e-cigarettes | 25 |
| HbA_1_C ≥10.0 gm% | 0 | Current smoker | 0 |
| **Body Mass Index** |  | **Sleep**  **Self-Reported Average Hours of Sleep per Night** |  |
|  |  | 7- <9 hours | 100 |
| <25.0 kg/m^2^ | 100 | 9- <10 hours | 90 |
| 25-29.9 kg/m^2^ | 70 | 6- <7 hours | 70 |
| 30-34.9 kg/m^2^ | 30 | 5- <6 or ≥10 hours | 40 |
| 35-39.9 kg/m^2^ | 15 | 4- <5 hours | 20 |
| ≥40 kg/m^2^ | 0 | <4 hours | 0 |

*20 points are subtracted if treated level**.**
